# Differential associations between neocortical tau pathology and blood flow with cognitive deficits in early-onset vs late-onset Alzheimer’s disease

**DOI:** 10.1101/2021.08.17.21262157

**Authors:** Denise Visser, Sander CJ Verfaillie, Emma E Wolters, Emma M Coomans, Tessa Timmers, Hayel Tuncel, Ronald Boellaard, Sandeep SV Golla, Albert D Windhorst, Philip Scheltens, Wiesje M van der Flier, Bart NM van Berckel, Rik Ossenkoppele

**Affiliations:** Department of Radiology & Nuclear Medicine, Amsterdam Neuroscience, Vrije Universiteit Amsterdam, Amsterdam UMC, Amsterdam, The Netherlands; Alzheimer Center Amsterdam,’ Department of Neurology, Amsterdam Neuroscience, Vrije Universiteit Amsterdam, Amsterdam UMC, Amsterdam, The Netherlands; Clinical Memory Research Unit, Lund University, Lund, Sweden; Department of Epidemiology and Biostatistics, Vrije Universiteit Amsterdam, Amsterdam UMC, Amsterdam, The Netherlands

**Keywords:** early-onset, Alzheimer’s disease, [^18^F]flortaucipir, tau pathology, cerebral blood flow, cognition

## Abstract

**Purpose:** Early-onset Alzheimer’s disease (EOAD) and late-onset Alzheimer’s disease (LOAD) differ in neuropathological burden and type of cognitive deficits. Assessing tau pathology and relative cerebral blood flow (rCBF) measured with [^18^F]flortaucipir PET in relation to cognition may help explain these differences between EOAD and LOAD.

**Methods:** Seventy-nine amyloid-positive individuals with a clinical diagnosis of AD (EOAD: n=35, age-at-PET=59±5, MMSE=23±4; LOAD: n=44, age-at-PET=71±5, MMSE=23±4) underwent a 130 minutes dynamic [^18^F]flortaucipir PET scan and extensive neuropsychological assessment. We extracted binding potentials (BP_ND_) and R_1_ (proxy of rCBF) from parametric images using receptor parametric mapping, in medial and lateral temporal, parietal, occipital and frontal regions-of-interest and used nine neuropsychological tests covering memory, attention, language and executive functioning. We first examined differences between EOAD and LOAD in BP_ND_ or R_1_ using ANOVA (region-of-interest analysis) and voxel-wise contrasts. Next, we performed linear regression models to test for potential interaction effects between age-at-onset and BP_ND_/R_1_ on cognition.

**Results:** Both region-of-interest and voxel-wise contrasts showed higher [^18^F]flortaucipir BP_ND_ values across all neocortical regions in EOAD. By contrast, LOAD patients had lower R_1_ values (indicative of more reduced rCBF) in medial temporal regions. For both tau and flow in lateral temporal, and occipito-parietal regions, associations with cognitive impairment were stronger in EOAD than in LOAD (EOAD BP_ND_ -0.76≤stβ≤-0.48 vs LOAD -0.18≤stβ≤-0.02; EOAD R_1_ 0.37≤stβ≤0.84 vs LOAD -0.25≤stβ≤0.16).

**Conclusions:** Compared to LOAD, the degree of lateral temporal and occipito-parietal tau pathology and relative cerebral blood-flow is more strongly associated with cognition in EOAD.

## Introduction

Alzheimer’s disease (AD) is characterized by depositions of amyloid-β plaques and hyperphosphorylated tau tangles[1]. The symptoms of AD are heterogeneous, partly related to age-at-onset. Late-onset AD is primarily associated with amnestic problems, while non-amnestic symptoms are more prominent when the disease manifests at younger age[2-6]. Atypical cognitive profiles in early-onset AD patients have previously been linked to specific spatial patterns of hypometabolism, atrophy, and more recently, tau pathology, that primarily affect the neocortex with relative sparing of the medial temporal lobe[7-10].

Age-at-onset is thus closely related to both cognitive symptoms and distinct patterns of tau pathology in AD. However, despite the notion that tau pathology is tightly linked to the degree of cognitive impairment in AD[11-17], it is currently unknown whether the association between tau pathology and cognitive performance is affected by age-at-onset. To address this question, we conducted a dynamic [^18^F]flortaucipir PET study. The dynamic scan protocol additionally yields a measure of R_1_ (a proxy of relative cerebral blood flow [rCBF][18]). In a previous study, we showed that [^18^F]flortaucipir R_1_ is strongly associated with cognitive performance in AD, (partly) independently of tau pathology[17]. We therefore also investigated differences in rCBF and its association with cognitive performance, taking into account age-at-onset.

Accordingly, we aimed to investigate the differences between early-onset AD and late-onset AD in 1) tau pathology and rCBF, and 2) the associations between tau pathology and rCBF with cognitive performance. We hypothesized that, relative to late-onset AD, early-onset AD displays higher levels of tau pathology. As R_1_ is tightly linked to hypometabolism measured with FDG-PET[19, 20], and lower levels of metabolism have previously been reported in early-onset AD patients, we expected to find lower levels of R_1_ in early-vs late-onset AD. Furthermore, as early-onset AD may represent a more ‘pure’ form of AD with relatively few comorbidities compared to late-onset AD[21], we hypothesized that tau pathology and cognitive performance would be more strongly associated in early-vs late-onset AD.

## Methods

### Participants

We included 79 subjects from the Amsterdam Dementia Cohort with either probable AD dementia[22] (n=68) or mild cognitive impairment (MCI) due-to-AD[23] (n = 11), with positive amyloid-β biomarkers[24-26]. All subjects underwent a standardized dementia screening, including medical history, extensive neuropsychological assessment, physical and neurological examination, lumbar puncture, blood tests, electroencephalography, and brain MRI. Diagnosis was established by consensus in a multidisciplinary meeting. The diagnosis of MCI/AD met core clinical criteria[22, 23] according to the National Institute on Aging and Alzheimer’s Association (NIA-AA) and were biomarker supported, with an AD-like CSF profile (i.e., tau/Aβ42 fraction >0.52)[27, 28] and/or an Aß-PET scan ([^11^C]PiB or [^18^F]florbetaben) showing substantial amyloid accumulation when visually assessed by an experienced nuclear medicine physician (BvB). When both CSF and PET measures of amyloid were available, PET visual read was used to determine amyloid status. Subjects were assigned to either the early- or late-onset AD group, based on a median split (age-at-PET). Since the median age in our sample was 66 years, this led to a similar age cut-off compared to the conventional threshold of 65 years[2, 3, 6, 29]. The sample included eight patients meeting criteria for an atypical variant of AD, including two behavioral variant AD (bvAD) and six posterior cortical atrophy (PCA) patients[26, 30]. All atypical AD patients were part of the early-onset AD group, except one bvAD patient. Exclusion criteria for all participants were (1) dementia not due-to-AD, (2) significant cerebrovascular disease on MRI (e.g. major cerebrovascular accident), (3) major traumatic brain injury, (4) major psychiatric or neurological disorders other than AD, (5) (recent) substance abuse. The study protocol was approved by the Medical Ethics Review Committee of the Amsterdam UMC, location VU Medical center. All patients provided written informed consent.

### [^18^F]flortaucipir PET

Acquisition and processing of [^18^F]flortaucipir PET images is described in detail elsewhere[17, 31, 32]. In short, dynamic 130min [^18^F]flortaucipir PET scans were acquired on a Philips Ingenuity TF-64 PET/CT scanner. The scanning protocol consisted of two dynamic PET scans of 60 and 50min, respectively, with an in-between 20min break[32]. The first 60min dynamic scan started simultaneously with a bolus injection of approximately 234±14 MBq [^18^F]flortaucipir (injected mass 1.2±0.9 μg). The second PET scan was co-registered to the first dynamic PET scan using Vinci software (Max Planck Institute, Germany). PET data were acquired in list mode and subsequently reconstructed using 3D-RAMLA including standard corrections for dead time, decay, attenuation, random, and scatter. Patients also underwent 3DT1-weighted MRI (Ingenuity TF PET/MR, Philips Medical Systems, The Netherlands) for anatomical and tissue segmentation purposes.

### Image analysis

The 3DT1-weighted MRI images were co-registered to their corresponding PET images using Vinci software. Anatomical regions-of-interest according to the Hammers template were subsequently delineated on the MR images and superimposed on the PET scan using PVElab[33]. Using receptor parametric mapping (RPM) with cerebellar gray matter as reference region we generated binding potential (BP_ND_) maps as a measure for tau pathology. Additionally, and similar to previous work[17, 31], R_1_ was generated as a proxy for rCBF[18]. In line with a previous study[17], we calculated BP_ND_ and R_1_ in frontal, occipital, parietal, medial and lateral temporal bilateral cortical lobar regions-of-interest.

In addition to the region-of-interest analyses, we performed voxel-wise analyses to 1) create average images of the different diagnostic groups (early- and late-onset AD without atypical AD cases, PCA and bvAD), and 2) explore more fine-grained differences in [^18^F]flortaucipir BP_ND_ and R_1_ between early- and late-onset AD. Therefore, we warped all native space parametric BP_ND_ and R_1_ images to Montreal Neurological Institute (MNI152) space using the transformation matrixes derived from warping the co-registered T1-weighted MRI scans to MNI space using Statistical Parametric Mapping (SPM) version 12. All warped images were visually inspected for transformation errors and quality control. Average images were calculated using SPM12.

### Neuropsychological assessment

We included eight neuropsychological tests (<1 year of PET scan), including the Dutch version of the Rey Auditory Verbal Learning Test immediate recall and delayed recall (episodic memory), Digit Span forward and backward (attention/executive functioning), Trail Making Test [TMT] version A and B (attention/executive functioning), Letter Fluency test (D-A-T) (executive functioning) and Category Fluency version animals (language)[34]. TMT-A and -B scores were inverted so that lower scores indicated worse performance. In addition, the Mini-Mental State Examination (MMSE) served as a measure of global cognition[35]. For participants who had TMT-A available but were missing TMT-B, we estimated the TMT-B by multiplying the time needed to complete the TMT-A with the mean TMT-B/A ratio from the respective diagnostic group in the respective cohort. To be included in the cognition analyses, participants were required to have at least 6 out of 9 cognitive tests available and 72 (91%) participants met this criterion (30 early- and 42 late-onset AD). Among the excluded cases there was one PCA case (early-onset AD group). sFigure-1 shows the missing values in the cognition-subsample for each neuropsychological test. There were on average 5% missing data in this subsample, for which we performed single imputation (with 5 iterations) for missing data, using all demographic (except for age and APOE4 status), neuropsychological and neuroimaging variables as predictors[36, 37].

### Statistical analysis

Differences in demographics and clinical characteristics between groups were assessed using independent sample t-tests for continuous variables and χ2 for dichotomous data. Differences in neuropsychological test-scores were assessed using ANOVA, adjusting for sex and education.

To assess differences in [^18^F]flortaucipir BP_ND_ and R_1_ between early- and late-onset AD groups, we used ANOVA with each of the lobar regions-of-interest (separately) as dependent variable. We repeated these analyses, but now excluding the atypical AD cases (n=8), to investigate whether potential age-dependent results were not driven solely by these extreme phenotypes. To assess more fine-grained differences, we additionally performed voxel-wise contrasts in SPM12 for BP_ND_ or R_1_ between early- and late-onset AD. All analyses were adjusted for sex. Results from voxel-wise analyses are displayed at both more liberal (i.e., p<0.001, uncorrected) and more stringent (p<0.05, Family Wise Error (FWE) corrected) thresholds. Next, we investigated whether associations between BP_ND_ or R_1_ and cognition differed between early- and late-onset AD. We performed linear regression analyses between BP_ND_ or R_1_ and neuropsychological tests (separate models), adjusted for sex and education, including the interaction term “age-at-onset (dichotomous)*BP_ND_ or R_1_”.

Results were considered significant if p-values were ≤0.05 for regional analyses. We considered a p-value ≤0.10 significant for interaction terms[38]. Additionally, adjustment for multiple comparisons was performed using the Benjamini-Hochberg False Discovery Rate (FDR) method (indicated by p_FDR_)[39]. Statistical analyses were performed using R software, version 4.0.2, and the ‘mice’ package was used for imputation.

## Results

Participant characteristics are presented in Table-1. There were no differences in level of education, sex distribution and APOE ε4 carriership between early- and late-onset AD (all p>0.05), also in the cognition subsample (sTable-1). Figure-1 and sFigure-2 show average BP_ND_ and R_1_ maps, respectively, to provide insight into their spatial distribution. On average, early-onset AD patients showed highest BP_ND_ values in medial and lateral parietal, and lateral temporal cortical areas (Fig-1). Late-onset AD patients showed highest BP_ND_ values in lateral temporal regions, and PCA patients in parieto-occipital regions, with BP_ND_ values exceeding all groups (Fig-1). Lastly, bvAD patients showed highest BP_ND_ values in lateral frontal and temporal regions, as well as lateral and medial parietal regions (Fig-1). For R_1_, values and patterns on the parametric images were visually similar across groups. (sFig-2).

**Table-1.** Demographics and cognitive test scores of the sample. Depicted are mean (SD), unless specified otherwise, for early-onset AD (EOAD) and late-onset AD (LOAD) groups. Median [range] is depicted for education. APOE ε4 status was unknown for four LOAD subjects. Independent sample T-test or χ2 test was used for demographic variables. Differences in cognition were assessed using ANOVA, adjusted for sex and education. *The Dutch Verhage scale for education includes 7 ascending categories, ranging from one (representing less than six years of primary education) to 7 (representing a university degree).

|  | **EOAD** | **LOAD** | **p-value** |
| --- | --- | --- | --- |
| **Sample size, n** | 35 | 44 |  |
| **Age-at-PET, y** | 59 (5) | 71 (5) | <0.001 |
| **Females, n (%)** | 19 (54) | 23 (52) | 0.724 |
| **APOE ε4 carriership, n** | 24/35 | 31/40 | 0.541 |
| **Education (Verhage scale\*)** | 6 [3-7] | 6 [3-7] | 0.318 |
| **MMSE** | 23 (4) | 23 (4) | 0.815 |
| **Cognition** |  |  |  |
| **Sample size (n)** | 30 | 42 |  |
| **RAVLT immediate recall** | 29 (8) | 23 (9) | 0.013 |
| **RAVLT delayed** | 3 (3) | 2 (3) | 0.329 |
| **Digit span forward** | 11 (2) | 11 (3) | 0.817 |
| **Digit span backward** | 7 (2) | 7 (3) | 0.069 |
| **Trail Making Test-A, sec** | 100 (88) | 68 (43) | 0.045 |
| **Trail Making Test-B, sec** | 334 (323) | 244 (167) | 0.112 |
| **Letter fluency test (D-A-T)** | 33 (12) | 32 (11) | 0.904 |
| **Category fluency Animals** | 17 (6) | 14 (5) | 0.076 |

**Figure-1.**
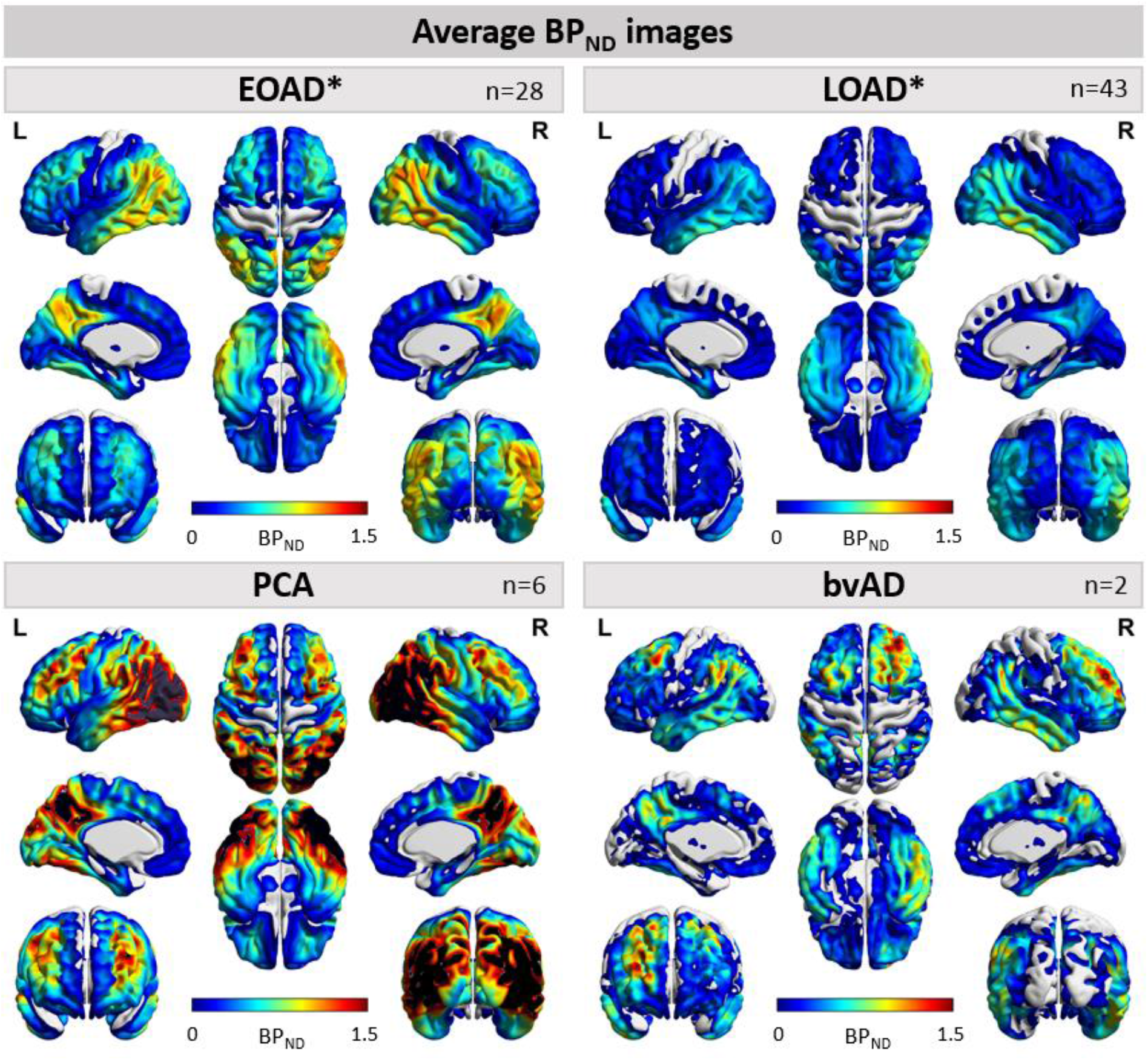
Average [^18^F]flortaucipir BP_ND_ images for early- and late-onset AD, PCA and bvAD. Average images of all early-onset Alzheimer’s disease (EOAD), late-onset AD (LOAD), posterior cortical atrophy (PCA) patients and behavioral variant AD (bvAD) patients on a scale ranging from BP_ND_ 0 to 1.5. * Excluding atypical variants, posterior cortical atrophy (PCA) patients and behavioral variant AD (bvAD) patients.

### Early- and late-onset AD differences in BP_ND_

Early-onset AD patients showed higher [^18^F]flortaucipir BP_ND_ in lateral temporal (BP_ND_ 0.56±0.30 vs 0.42±0.30, p=0.045), parietal (BP_ND_ 0.84±0.50 vs 0.33±0.29, p<0.001), occipital (BP_ND_ 0.65±0.52 vs 0.29±0.23, p<0.001) and frontal cortex (BP_ND_ 0.40±0.30 vs 0.16±0.23, p<0.001) compared to late-onset AD (Fig-2).There were no differences in medial temporal BP_ND_ between early-onset AD (0.24±0.14) and late-onset AD (0.25±0.18, p=0.895). Voxel-wise analyses confirmed higher BP_ND_ in widespread neocortical regions in early-vs late-onset AD (Fig-2G). Effects were most pronounced in the precuneus and posterior cingulate, and fronto-temporal cortex, as supported by the FWE-corrected results. There were no regions in which late-onset AD showed higher BP_ND_ compared to early-onset AD (Fig-2G). Results remained essentially unchanged when PCA and bvAD patients were excluded from the analysis (sFig-3).

**Figure-2.**
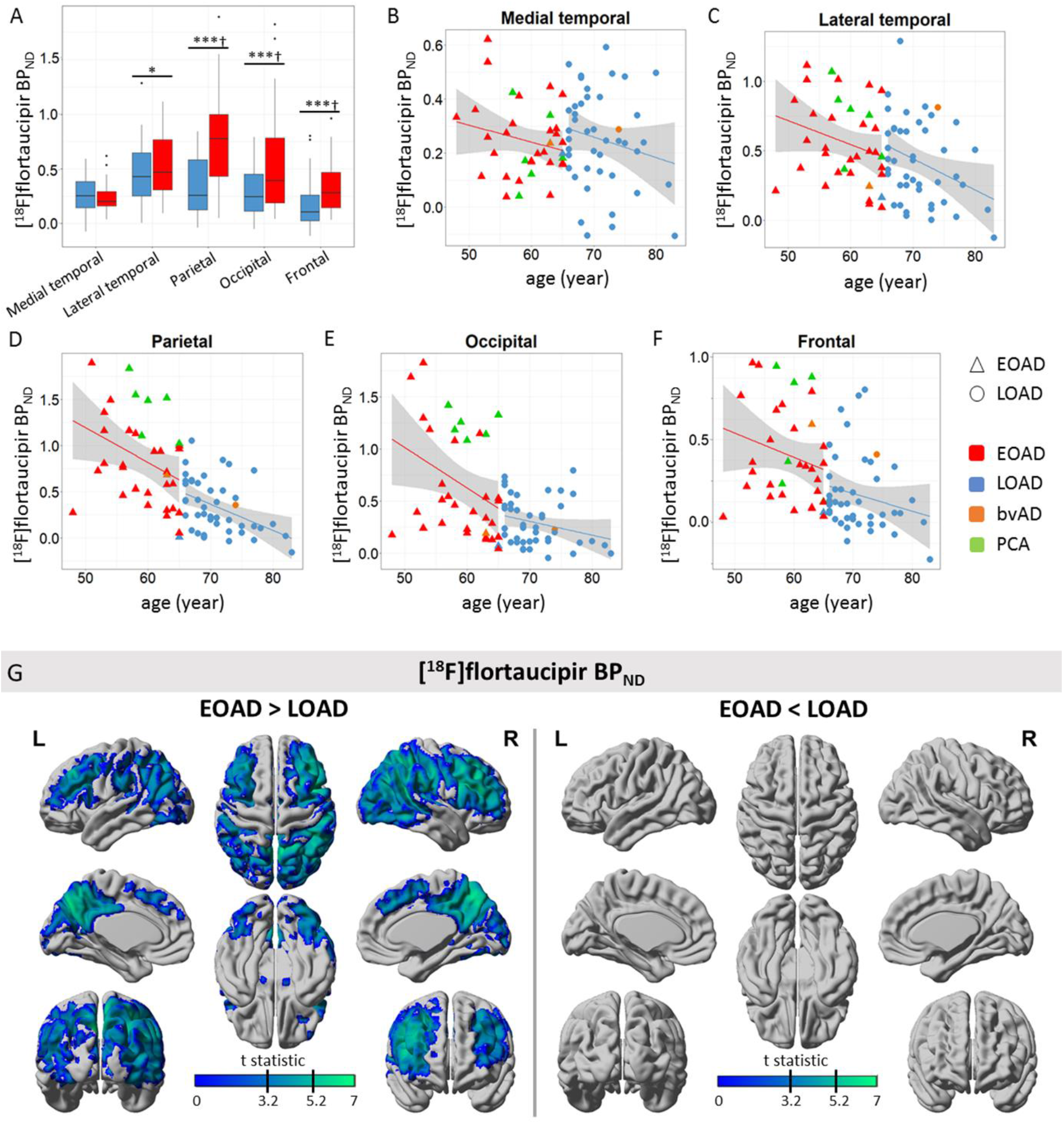
[^18^F]flortaucipir BP_ND_ for early- and late-onset AD. A) Boxplot of [^18^F]flortaucipir BP_ND_ values for each region-of-interest (ROI). Differences were assessed using ANOVA, adjusted for sex. B-F) Scatterplots for [^18^F]flortaucipir BP_ND_ by age for each ROI. G) Results from voxel-wise contrast for [^18^F]flortaucipir BP_ND_ between early-onset AD (EOAD) and late-onset AD (LOAD). Indicated by the black lines on the color scale are thresholds for p<0.001, uncorrected (t = 3.20) and for p<0.05, FWE-corrected (t = 5.24). *p<0.05, ^**^p<0.01, ***p<0.001, †p_FDR_<0.05.

### Early-and late-onset AD differences in R_1_

By contrast, [^18^F]flortaucipir R_1_ was lower in late-onset AD compared to early-onset AD in the medial temporal lobe (R_1_ 0.66±0.05 vs 0.69±0.05, p<0.05), but not in any of the neocortical regions (p>0.05) (Fig-3). Voxel-wise analyses showed lower R_1_ in the (medial) temporal lobe (surviving FWE-correction), and subtly lower R_1_ in the medial frontal cortex in late-onset compared to early-onset AD (Fig-3G). In contrast, parieto-occipital regions showed lower R_1_ in early-onset AD compared to late-onset AD, but this did not survive FWE-correction. Results remained essentially unchanged when atypical cases were excluded from the analysis (sFig-4).

**Figure-3.**
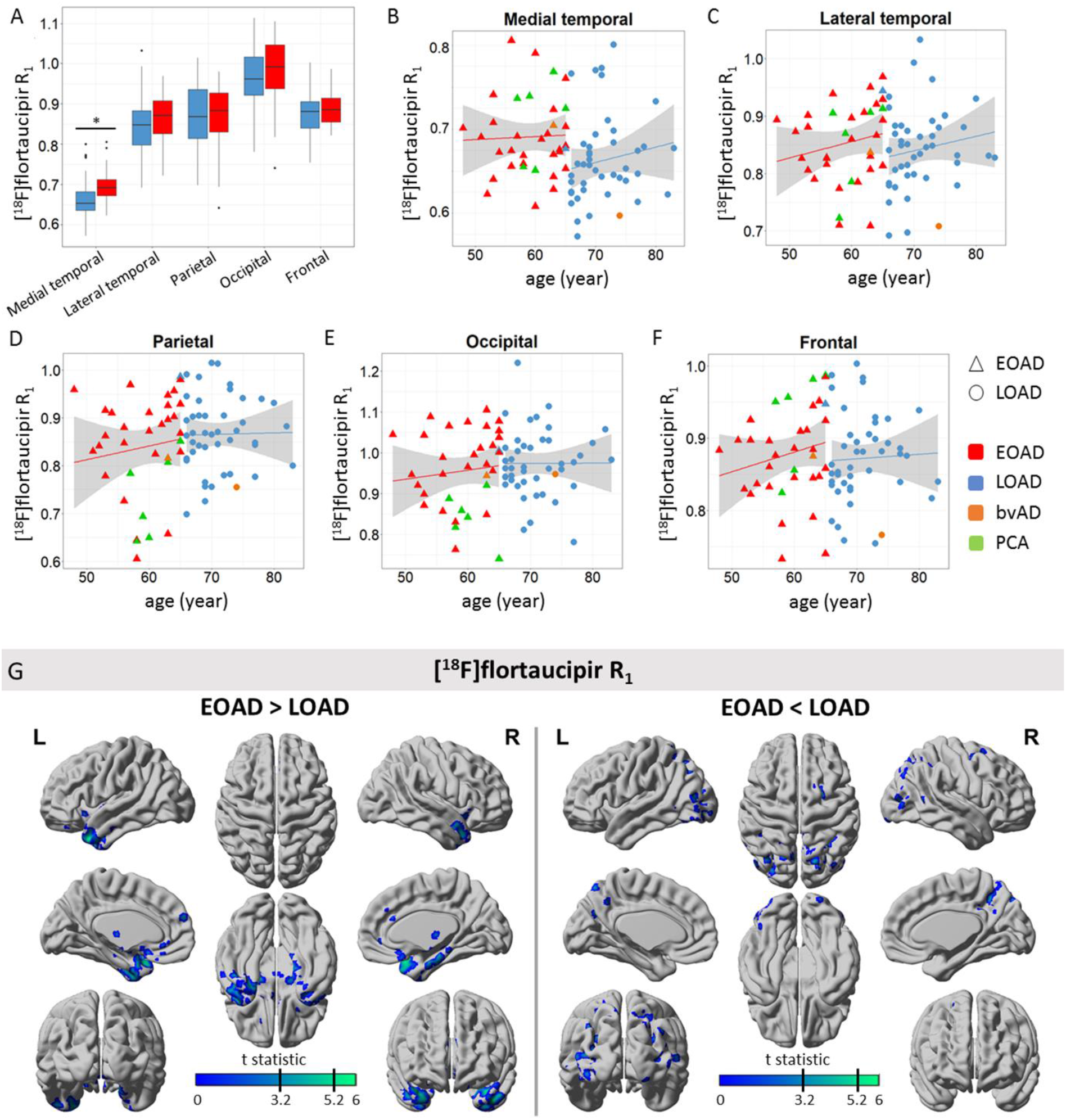
[^18^F]flortaucipir R_1_ for early- and late-onset AD. A) Boxplot of [^18^F]flortaucipir R_1_ values for each region-of-interest (ROI). B-F) Scatterplots for [^18^F]flortaucipir R_1_ by age for each ROI. G) Results from voxel-wise contrast for [^18^F]flortaucipir R_1_ between early-onset AD (EOAD) and late-onset AD (LOAD). Indicated by the black lines on the color scale are thresholds for p<0.001, uncorrected (t = 3.20) and for p<0.05, FWE-corrected (t = 5.24). *p<0.05.

### Early- and late-onset AD differences in the association between BP_ND_ and cognition

Linear regression analyses showed that, in general, higher [^18^F]flortaucipir BP_ND_ was strongly associated with worse scores on a variety of cognitive tests (Fig-4A). Age-at-onset moderated these associations such that in early-onset AD, associations between lateral temporal, parietal and occipital BP_ND_ with TMT-A, and parietal BP_ND_ with TMT-B were stronger than in late-onset AD (all p_interaction_<0.10; Fig-4). In late-onset AD, associations between medial temporal BP_ND_ and Digit Span backward and Letter fluency test (D-A-T) were stronger than in early-onset AD (Fig-4).

**Figure-4.**
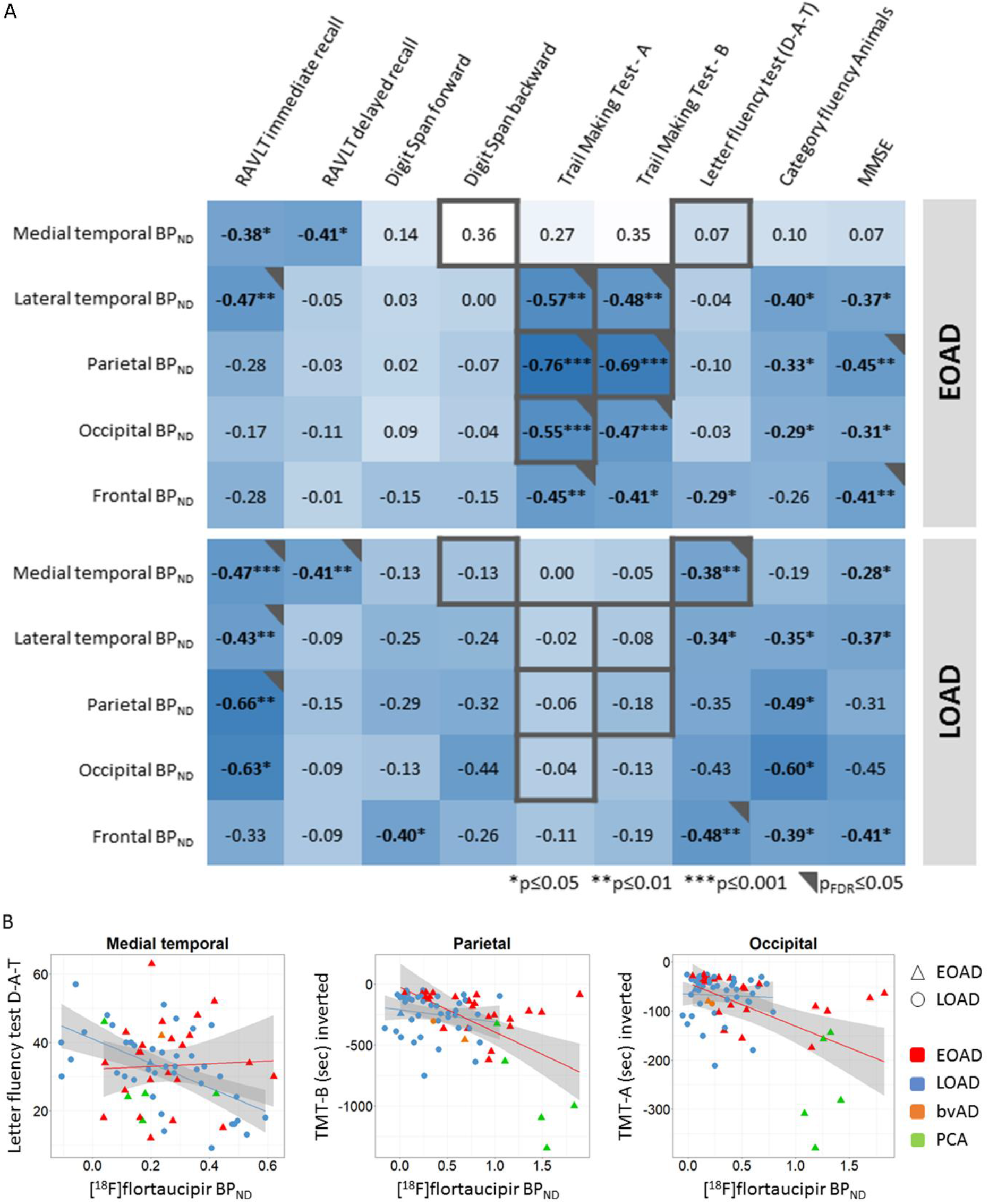
Associations between [^18^F]flortaucipir BP_ND_ and cognitive test scores for early- and late-onset AD. A) Significant modification of age-at-onset as assessed in the model including all AD subjects from the cognition subsample (n=72), adjusted for age, sex, and education is depicted in grey (representing interaction terms at p≤0.10) and black (representing interaction terms at p_FDR_≤0.05) squares. Standardized regression coefficients are depicted for early-onset AD (EOAD) and late-onset AD (LOAD) separately. B) A selection of scatterplots for the association between [^18^F]flortaucipir BP_ND_ and neuropsychological test scores.

### Early- and late-onset AD differences in the association between R_1_ and cognition

In general, lower [^18^F]flortaucipir R_1_ was associated with worse scores on multiple cognitive tests (Fig-5). Age-at-onset moderated these associations such that in early-onset AD, associations were stronger than in late-onset AD (all p_interaction_<0.10; Fig-5). More specifically, this was the case for the association between lateral temporal R_1_ and Digit Span backward, TMT-A, TMT-B, and Letter fluency test (D-A-T), and the associations between occipito-parietal R_1_ and TMT-A, TMT-B, and Category fluency Animals, as well as the association between occipital R_1_ and MMSE (Fig-5).

**Figure-5.**
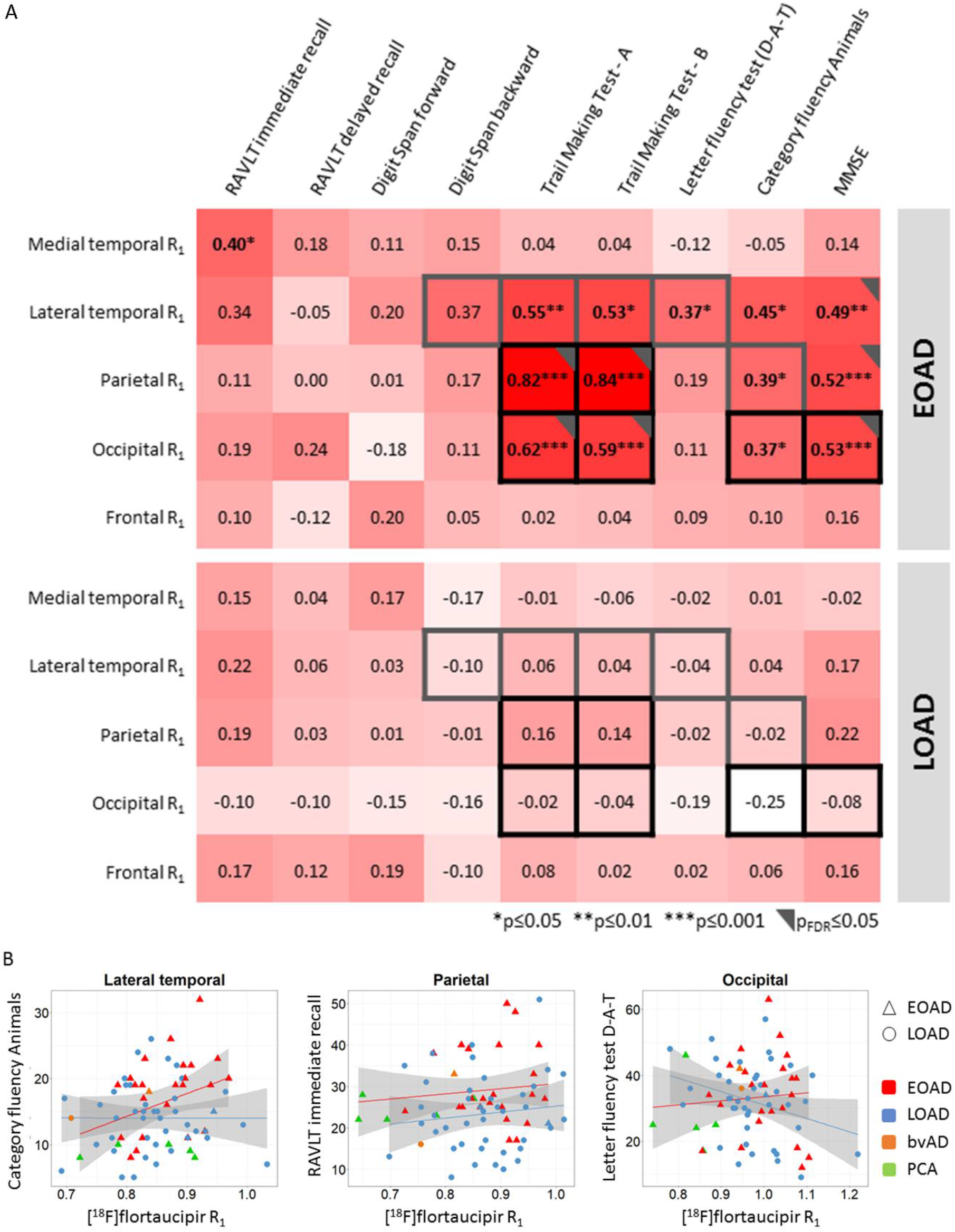
Associations between [^18^F]flortaucipir R_1_ and cognitive test scores for early- and late-onset AD. A) Significant modification of age-at-onset as assessed in the model including all AD subjects from the cognition subsample (n=72), adjusted for age, sex, and education is depicted in grey (representing interaction terms at p≤0.10) and black (representing interaction terms at p_FDR_≤0.05) squares. Standardized regression coefficients are depicted for early- and late-onset AD separately. B) Selection of scatterplots for the association between [^18^F]flortaucipir R_1_ and neuropsychological test scores.

## Discussion

The main findings of this study include higher levels of neocortical tau pathology in early-onset AD compared to late-onset AD, while late-onset AD showed reduced rCBF in the medial temporal lobe compared to early-onset AD. Moreover, we found that higher levels of tau pathology and lower cerebral blood flow in lateral temporal, and occipito-parietal regions were more strongly associated with cognitive impairment (mainly executive functioning domain) in early-vs late-onset AD. Previous studies demonstrated that, relative to late-onset AD, early-onset AD exhibits more extensive pathological and neurodegenerative changes with respect to (among others) amyloid, metabolic activity[7, 40], atrophy[41, 42], functional network changes[43-45] and tau[46, 47]. The current study extends on these previous studies not only showing greater tau load in early-vs late-onset AD, but also providing novel insights into age-dependent differences in cerebral blood flow, and demonstrates differential associations of both biomarkers with cognition. These findings contribute to our understanding of the differences between early- and late-onset AD patients and may support that younger AD patients are more suitable for inclusion in clinical trials, as the stronger link between pathology and cognitive decline suggests that a greater benefit may be achieved in this population when targeting tau pathology or cerebral blood flow compared to older AD patients, where this link is less strong. Besides, our results emphasize the importance of cerebrovascular health (and its potential treatment) in younger AD patients specifically.

For tau pathology, associations with worse cognitive performance across non-memory domains were stronger in early-onset AD, apart from associations in the medial temporal lobe. The medial temporal region is predominantly involved in sporadic (late-onset) AD[16, 45, 48], and in line with this, associations with cognitive performance for this region were stronger in late-onset AD. For rCBF, we found that lower blood flow was associated with a higher degree of cognitive impairment in non-memory domains in early-vs late-onset AD. This indicates that higher levels of neocortical tau pathology and lower rCBF have a relatively stronger influence on cognitive (dys)functioning in early-onset AD. The main hypothesis for these findings is that co-pathologies (e.g. other proteinopathies such as TDP-43 or alpha-synuclein, or vascular damage) often developing at older age, may contribute to progressive cognitive impairment in late-onset AD[49-51]. In line with this, a recent study showed that the prevalence and severity of most comorbid pathologies was lower in early-vs late-onset AD[21]. Another explanation is that the brain regions most heavily affected in early-onset AD, are more important for broader cognitive functioning (other than memory-specific), and comprise specific neuronal networks crucial for specific cognitive functions. The parietal lobe contains a high pathologic burden in early-onset AD (as shown by the result of the current study and by others (e.g.[7, 45, 47])), and is an important hub in higher-order cognitive networks like the executive-control network[52, 53]. If networks like these, or their cross-network relationships, segregate with older age[54] it might be that the relative importance of the regions affected by either tau pathology or decreases in rCBF decrease with older age. This would be reflected by a weakened association between tau pathology or rCBF and cognition (executive functioning specifically) in late-compared to early-onset AD.

In the current study we used R_1_ images as a measure of cerebral blood flow. This measure is tightly linked to hypometabolism measured with FDG-PET[19, 20], and others have shown decreased occipito-parietal glucose metabolism in early-onset AD[7, 55]. In contrast, our regional analyses did not show lower rCBF in early-vs late-onset AD. However, additional voxel-wise comparisons showed more fine grained decreased rCBF in lateral parietal and occipital regions in early-onset AD. One explanation for these findings might be that there are no extensive or clear differential patterns of cortical reductions in rCBF in early-vs late-onset AD, or that differential reductions in rCBF may be restricted to specific cortical gyri. Another explanation could be that R_1_, which is a proxy of rCBF, is not sensitive enough to capture the differences between early- and late-onset AD. As rCBF decreases with age[56], this might suggest that early-onset AD patients are more severely affected by deficits in rCBF compared to late-onset AD patients, which in turn might be caused (in part) by the higher levels of tau pathology present. Ideally one would investigate whether early-onset AD patients are indeed more heavily affected by reduced rCBF by including age-matched control groups, but since we had no such data available, it remains to be elucidated in future research.

Our results showed that early-onset AD patients exhibit higher levels of tau pathology in widespread neocortical regions compared to late-onset AD patients, but no differences were found in the medial temporal lobe. Others found higher levels of tau pathology primarily in (pre)frontal and (inferior) parietal cortices in early-relative to late-onset AD, and no differences in the medial temporal cortex[47]. Another study similarly showed that early-onset AD patients, showed greater binding in the inferior parietal, occipital, and inferior temporal cortices[57]. The pattern of cortical involvement as found in the current study thus are highly consistent with previous findings, indicating that the development of high levels of tau pathology follows a specific spatial pattern within early-onset AD patients. Supporting this, a recent study identified four distinct spatiotemporal trajectories of tau pathology in AD, each presenting with distinct demographic and cognitive profiles and differing longitudinal outcomes. The medial temporal lobe-sparing subtype for example, was associated with younger age, less APOE4 allele carriership, and greater overall tau burden[58], which is largely in accordance with findings in our early-onset AD study population. Others point towards genetics explaining higher tau burden, since comparable results were found in autosomal dominant mutation carriers (e.g. presenilin-1)[59, 60]. The differential spatial patterns of tau pathology in early-onset AD might also be explained by genetic involvement, as early-onset AD patients are less frequently APOE4 allele carriers, and E4 genotype does influence spatial patterns of brain pathology[61-63]. In the present study, however, we did find slightly, but non-significantly lower E4 allele carriership in early-onset AD, which makes it unlikely that E4 genotype may have explained the findings.

A strength of this study is usage of a single dynamic [^18^F]flortaucipir PET scan to derive measures of both tau pathology (BP_ND_) and relative cerebral blood flow (R_1_). There were also several limitations. First, the late-onset AD patients in our study were relatively young, which might hamper generalizability of results to older study populations. Second, seven participants did not have sufficient neuropsychological data available for cognition analyses. Although results are not expected to be influenced to a large extent, it might be that results as described in this study are slightly underestimating the differential effect of age-at-onset, given that neuropsychological data were missing in the most severely progressed patients (mean MMSE score: 20±4). Third, the cross-sectional design did not allow investigating the temporal evolution of tau pathology, rCBF and cognitive impairment. Future studies with longitudinal data available could aid in elucidating the timely pathways of these three parameters.

In conclusion, early-onset AD is characterized by higher levels of tau pathology and stronger associations between lateral temporal, and occipito-parietal tau pathology or lower rCBF and cognitive impairment. These findings may have important implications for clinical trials, since effects of potential tau- or blood flow targeting therapeutic interventions might exert larger effects in early-onset AD compared to late-onset AD patients. Furthermore, differences between early- and late-onset AD patients as described in the current study should be considered in any therapeutic intervention trial were either cognition is used as an outcome measure or where indirect effects on tau pathology or rCBF are expected, to ensure correct interpretation of results and aid in the formation of appropriate patient selection criteria.

## Data Availability

Anonymized data used in the present study may be available upon request to the corresponding author.

## Compliance with Ethical Standards

### Funding

This study was funded by a ZonMW Memorabel grant.

### Conflict of Interest

Visser, Verfaillie, Wolters, Coomans, Timmers, Tuncel, Boellaard, Golla, Windhorst and Ossenkoppele declares that he/she has no conflict of interest.

Van der Flier received grant support from ZonMW, NWO, EU-FP7, Alzheimer Nederland, CardioVascular Onderzoek Nederland, Stichting Dioraphte, Gieskes-Strijbis Fonds, Boehringer Ingelheim, Piramal Neuroimaging, Roche BV, Janssen Stellar, Combinostics. All funding is paid to the institution. WvdF holds the Pasman chair.

Van Berckel received research support from ZonMW, AVID radiopharmaceuticals, CTMM and Janssen Pharmaceuticals. He is a trainer for Piramal and GE. He receives no personal honoraria.

Scheltens received grant support (to the institution) from GE Healthcare, Danone Research, Piramal and MERCK. In the past 2 years he has received consultancy/speaker fees from Lilly, GE Healthcare, Novartis, Forum, Sanofi, Nutricia, Probiodrug and EIP Pharma. All funding is paid to the institution.

No other potential conflicts of interest relevant to this article exist.

### Ethical approval

All procedures performed in studies involving human participants were in accordance with the ethical standards of the institutional and/or national research committee and with the 1964 Helsinki declaration and its later amendments or comparable ethical standards.

### Informed consent

Informed consent was obtained from all individual participants included in the study.

### Disclaimer

Denise Visser and Sander CJ Verfaillie contributed equally to this work.

## Acknowledgments

We kindly thank all participants for their contribution. Research of Amsterdam Alzheimer Center is part of the Neurodegeneration program of Amsterdam Neuroscience. The Amsterdam Alzheimer Center is supported by Alzheimer Nederland and Stichting VUmc funds. [^18^F]Flortaucipir PET scans were made possible by Avid Radiopharmaceuticals Inc.

## Notes

### Author Declarations

The study protocol was approved by the Medical Ethics Review Committee of the Amsterdam UMC, location VU Medical center.

## References

1. Jack Jr, C.R., et al., NIA-AA research framework: toward a biological definition of Alzheimer’s disease. Alzheimer’s & Dementia, 2018. 14(4): p. 535–562.

2. Koedam, E.L., et al., Early-versus late-onset Alzheimer’s disease: more than age alone. Journal of Alzheimer’s Disease, 2010. 19(4): p. 1401–1408.

3. Koedam, E.L., et al., Early-onset dementia is associated with higher mortality. Dementia and geriatric cognitive disorders, 2008. 26(2): p. 147–152.

4. Koss, E., et al., Clinical and neuropsychological differences between patients with earlier and later onset of Alzheimer’s disease: A CERAD analysis, Part XII. Neurology, 1996. 46(1): p. 136–141.

5. Scheltens, N.M., et al., The identification of cognitive subtypes in Alzheimer’s disease dementia using latent class analysis. Journal of Neurology, Neurosurgery & Psychiatry, 2016. 87(3): p. 235–243.

6. Smits, L.L., et al., Early onset Alzheimer’s disease is associated with a distinct neuropsychological profile. Journal of Alzheimer’s Disease, 2012. 30(1): p. 101–108.

7. Ossenkoppele, R., et al., Amyloid burden and metabolic function in early-onset Alzheimer’s disease: parietal lobe involvement. Brain, 2012. 135(7): p. 2115–2125.

8. Smits, L.L., et al., Regional atrophy is associated with impairment in distinct cognitive domains in Alzheimer’s disease. Alzheimer’s & Dementia, 2014. 10: p. S299–S305.

9. Hsu, J.-L., et al., Posterior atrophy and medial temporal atrophy scores are associated with different symptoms in patients with Alzheimer’s disease and mild cognitive impairment. PloS one, 2015. 10(9): p. e0137121.

10. Whitwell, J.L., et al., Neuroimaging correlates of pathologically defined subtypes of Alzheimer’s disease: a case-control study. The Lancet Neurology, 2012. 11(10): p. 868–877.

11. Pontecorvo, M.J., et al., Relationships between flortaucipir PET tau binding and amyloid burden, clinical diagnosis, age and cognition. Brain, 2017. 140(3): p. 748–763.

12. Aschenbrenner, A.J., et al., Influence of tau PET, amyloid PET, and hippocampal volume on cognition in Alzheimer disease. Neurology, 2018. 91(9): p. e859–e866.

13. Johnson, K.A., et al., Tau positron emission tomographic imaging in aging and early Alzheimer disease. Annals of neurology, 2016. 79(1): p. 110–119.

14. Ossenkoppele, R., et al., Tau PET patterns mirror clinical and neuroanatomical variability in Alzheimer’s disease. Brain, 2016. 139(5): p. 1551–1567.

15. Ossenkoppele, R., et al., Associations between tau, Aβ, and cortical thickness with cognition in Alzheimer disease. Neurology, 2019. 92(6): p. e601–e612.

16. Schöll, M., et al., PET imaging of tau deposition in the aging human brain. Neuron, 2016. 89(5): p. 971–982.

17. Visser, D., et al., Tau pathology and relative cerebral blood flow are independently associated with cognition in Alzheimer’s disease. European journal of nuclear medicine and molecular imaging, 2020. 47: p. 3165–3175.

18. Golla, S.S., et al., Parametric methods for [18F] flortaucipir PET. Journal of Cerebral Blood Flow & Metabolism, 2020. 40(2): p. 365–373.

19. Peretti, D.E., et al., Relative cerebral flow from dynamic PIB scans as an alternative for FDG scans in Alzheimer’s disease PET studies. PloS one, 2019. 14(1): p. e0211000.

20. Ottoy, J., et al., 18F-FDG PET, the early phases and the delivery rate of 18F-AV45 PET as proxies of cerebral blood flow in Alzheimer’s disease: Validation against 15O-H2O PET. Alzheimer’s & Dementia, 2019. 15(9): p. 1172–1182.

21. Spina, S., et al., Comorbid neuropathological diagnoses in early versus late-onset Alzheimer’s disease. Brain, 2021.

22. McKhann, G.M., et al., The diagnosis of dementia due to Alzheimer’s disease: recommendations from the National Institute on Aging-Alzheimer’s Association workgroups on diagnostic guidelines for Alzheimer’s disease. Alzheimer’s & dementia, 2011. 7(3): p. 263–269.

23. Albert, M.S., et al., The diagnosis of mild cognitive impairment due to Alzheimer’s disease: recommendations from the National Institute on Aging-Alzheimer’s Association workgroups on diagnostic guidelines for Alzheimer’s disease. Focus, 2013. 11(1): p. 96–106.

24. van der Flier, W.M., et al., Optimizing patient care and research: the Amsterdam Dementia Cohort. Journal of Alzheimer’s disease, 2014. 41(1): p. 313–327.

25. Crutch, S.J., et al., Posterior cortical atrophy. The Lancet Neurology, 2012. 11(2): p. 170–178.

26. Ossenkoppele, R., et al., The behavioural/dysexecutive variant of Alzheimer’s disease: clinical, neuroimaging and pathological features. Brain, 2015. 138(9): p. 2732–2749.

27. Tijms, B.M., et al., Unbiased approach to counteract upward drift in cerebrospinal fluid amyloid-β 1–42 analysis results. Clinical Chemistry, 2018. 64(3): p. 576–585.

28. Duits, F.H., et al., The cerebrospinal fluid “Alzheimer profile”: easily said, but what does it mean? Alzheimer’s & Dementia, 2014. 10(6): p. 713-723. e2.

29. Mendez, M.F., et al., Nonamnestic presentations of early-onset Alzheimer’s disease. American Journal of Alzheimer’s Disease & Other Dementias®, 2012. 27(6): p. 413–420.

30. Crutch, S.J., et al., Consensus classification of posterior cortical atrophy. Alzheimer’s & Dementia, 2017. 13(8): p. 870–884.

31. Wolters, E.E., et al., Tau pathology, relative cerebral flow and cognition in dementia with Lewy bodies: Tau imaging. Alzheimer’s & Dementia, 2020. 16: p. e041048.

32. Golla, S.S., et al., Quantification of tau load using [18 F] AV1451 PET. Molecular imaging and biology, 2017. 19(6): p. 963–971.

33. Rask, T., et al., PVElab: Software for correction of functional images for partial volume errors. Neuroimage, 2004. 22.

34. Groot, C., et al., Differential effects of cognitive reserve and brain reserve on cognition in Alzheimer disease. Neurology, 2018. 90(2): p. e149–e156.

35. Folstein, M.F., S.E. Folstein, and P.R. McHugh, “Mini-mental state”: a practical method for grading the cognitive state of patients for the clinician. Journal of psychiatric research, 1975. 12(3): p. 189–198.

36. Rubin, D.B., Multiple imputation for nonresponse in surveys. Vol. 81. 2004: John Wiley & Sons.

37. Von Hippel, P.T., How many imputations do you need? A two-stage calculation using a quadratic rule. Sociological Methods & Research, 2020. 49(3): p. 699–718.

38. Selvin, S., Statistical analysis of epidemiologic data. 2004: Oxford University Press.

39. Benjamini, Y. and Y. Hochberg, Controlling the false discovery rate: a practical and powerful approach to multiple testing. Journal of the Royal statistical society: series B (Methodological), 1995. 57(1): p. 289–300.

40. Lehmann, M., et al., Diverging patterns of amyloid deposition and hypometabolism in clinical variants of probable Alzheimer’s disease. Brain, 2013. 136(3): p. 844–858.

41. Chan, D., et al., Change in rates of cerebral atrophy over time in early-onset Alzheimer’s disease: longitudinal MRI study. The Lancet, 2003. 362(9390): p. 1121–1122.

42. Möller, C., et al., Different patterns of gray matter atrophy in early-and late-onset Alzheimer’s disease. Neurobiology of aging, 2013. 34(8): p. 2014–2022.

43. Lehmann, M., et al., Loss of functional connectivity is greater outside the default mode network in nonfamilial early-onset Alzheimer’s disease variants. Neurobiology of aging, 2015. 36(10): p. 2678–2686.

44. De Waal, H., et al., EEG abnormalities in early and late onset Alzheimer’s disease: understanding heterogeneity. Journal of Neurology, Neurosurgery & Psychiatry, 2011. 82(1): p. 67–71.

45. De Waal, H., et al., Young Alzheimer patients show distinct regional changes of oscillatory brain dynamics. Neurobiology of aging, 2012. 33(5): p. 1008.e25-1008.e31.

46. Iaccarino, L., et al., Spatial Relationships between molecular pathology and neurodegeneration in the Alzheimer’s disease continuum. Cerebral Cortex, 2021. 31(1): p. 1–14.

47. Schöll, M., et al., Distinct 18F-AV-1451 tau PET retention patterns in early-and late-onset Alzheimer’s disease. Brain, 2017. 140(9): p. 2286–2294.

48. Cavedo, E., et al., Medial temporal atrophy in early and late-onset Alzheimer’s disease. Neurobiology of aging, 2014. 35(9): p. 2004–2012.

49. Ossenkoppele, R., et al., Accuracy of Tau Positron Emission Tomography as a Prognostic Marker in Preclinical and Prodromal Alzheimer Disease: A Head-to-Head Comparison Against Amyloid Positron Emission Tomography and Magnetic Resonance Imaging. JAMA neurology, 2021.

50. Gerritsen, A.A., et al., Prevalence of comorbidity in patients with young-onset Alzheimer disease compared with late-onset: a comparative cohort study. Journal of the American Medical Directors Association, 2016. 17(4): p. 318–323.

51. Ortner, M., et al., Small vessel disease, but neither amyloid load nor metabolic deficit, is dependent on age at onset in Alzheimer’s disease. Biological psychiatry, 2015. 77(8): p. 704–710.

52. Seeley, W.W., et al., Neurodegenerative diseases target large-scale human brain networks. Neuron, 2009. 62(1): p. 42–52.

53. Brier, M.R., et al., Loss of intranetwork and internetwork resting state functional connections with Alzheimer’s disease progression. Journal of Neuroscience, 2012. 32(26): p. 8890–8899.

54. Engels, M., et al., Alzheimer’s disease: the state of the art in resting-state magnetoencephalography. Clinical Neurophysiology, 2017. 128(8): p. 1426–1437.

55. Rabinovici, G.D., et al., Increased metabolic vulnerability in early-onset Alzheimer’s disease is not related to amyloid burden. Brain, 2010. 133(2): p. 512–528.

56. Chen, J.J., H.D. Rosas, and D.H. Salat, Age-associated reductions in cerebral blood flow are independent from regional atrophy. Neuroimage, 2011. 55(2): p. 468–478.

57. Cho, H., et al., Excessive tau accumulation in the parieto-occipital cortex characterizes earlyonset Alzheimer’s disease. Neurobiology of aging, 2017. 53: p. 103–111.

58. Vogel, J.W., et al., Four distinct trajectories of tau deposition identified in Alzheimer’s disease. Nature Medicine, 2021: p. 1–11.

59. Smith, R., et al., Posterior accumulation of tau and concordant hypometabolism in an earlyonset Alzheimer’s disease patient with presenilin-1 mutation. Journal of Alzheimer’s Disease, 2016. 51(2): p. 339–343.

60. Gordon, B.A., et al., Tau PET in autosomal dominant Alzheimer’s disease: relationship with cognition, dementia and other biomarkers. Brain, 2019. 142(4): p. 1063–1076.

61. van der Flier, W.M., et al., Early-onset versus late-onset Alzheimer’s disease: the case of the missing APOE ε4 allele. The Lancet Neurology, 2011. 10(3): p. 280–288.

62. Ten Kate, M., et al., Impact of APOE-ε4 and family history of dementia on gray matter atrophy in cognitively healthy middle-aged adults. Neurobiology of aging, 2016. 38: p. 14–20.

63. Mattsson, N., et al., Greater tau load and reduced cortical thickness in APOE ε4-negative Alzheimer’s disease: a cohort study. Alzheimer’s research & therapy, 2018. 10(1): p. 1–12.

