## Supplementary material for "Differential associations between neocortical tau pathology and blood flow with cognitive deficits in early-onset vs late-onset Alzheimer’s disease"

| | EOAD | LOAD | p-values |
|---|---|---|---|
| **Sample size (n)** | 30 | 42 | |
| **Age (years)** | 59 (5) | 71 (5) | <0.001 |
| **Females n (%)** | 14 (47) | 19 (45) | ~1 |
| **Education (Verhage scale*)** | 6 [3-7] | 5 [3-7] | 0.637 |
| **MMSE** | 23 (3) | 23 (4) | 0.708 |
| **APOE4 ε4 carriership, n/$n_{total}$** | 21/30 | 33/38 | 0.757 |
| **[18F]flortaucipir $BP_{ND}$** | | | |
| **Medial temporal** | 0.24 (0.14) | 0.25 (0.18) | 0.842 |
| **Lateral temporal** | 0.55 (0.31) | 0.42 (0.30) | 0.081 |
| **Parietal** | 0.84 (0.51) | 0.33 (0.29) | <0.001 |
| **Occipital** | 0.64 (0.54) | 0.29 (0.23) | <0.001 |
| **Frontal** | 0.38 (0.30) | 0.16 (0.23) | <0.001 |
| **[18F]flortaucipir $R_1$** | | | |
| **Medial temporal** | 0.69 (0.04) | 0.66 (0.05) | 0.004 |
| **Lateral temporal** | 0.86 (0.06) | 0.84 (0.07) | 0.155 |
| **Parietal** | 0.86 (0.09) | 0.87 (0.08) | 0.794 |
| **Occipital** | 0.97 (0.09) | 0.97 (0.08) | 0.891 |
| **Frontal** | 0.89 (0.06) | 0.87 (0.06) | 0.209 |

**sTable-1. Demographics, [18F]flortaucipir $BP_{ND}$ and $R_1$ of n=72 (cognition) sample.** Depicted are mean (SD), unless specified otherwise, for early-onset AD (EOAD) and late-onset AD (LOAD) groups. Median [range] is depicted for education. APOE ε4 status was unknown for four LOAD subjects. Independent sample T-test or $\chi 2$ test was used for demographic variables. Differences in [18F]flortaucipir $BP_{ND}$ or $R_1$ were assessed using ANOVA, adjusted for sex. *The Dutch Verhage scale for education includes 7 ascending categories, ranging from one (representing less than six years of primary education) to 7 (representing a university degree).

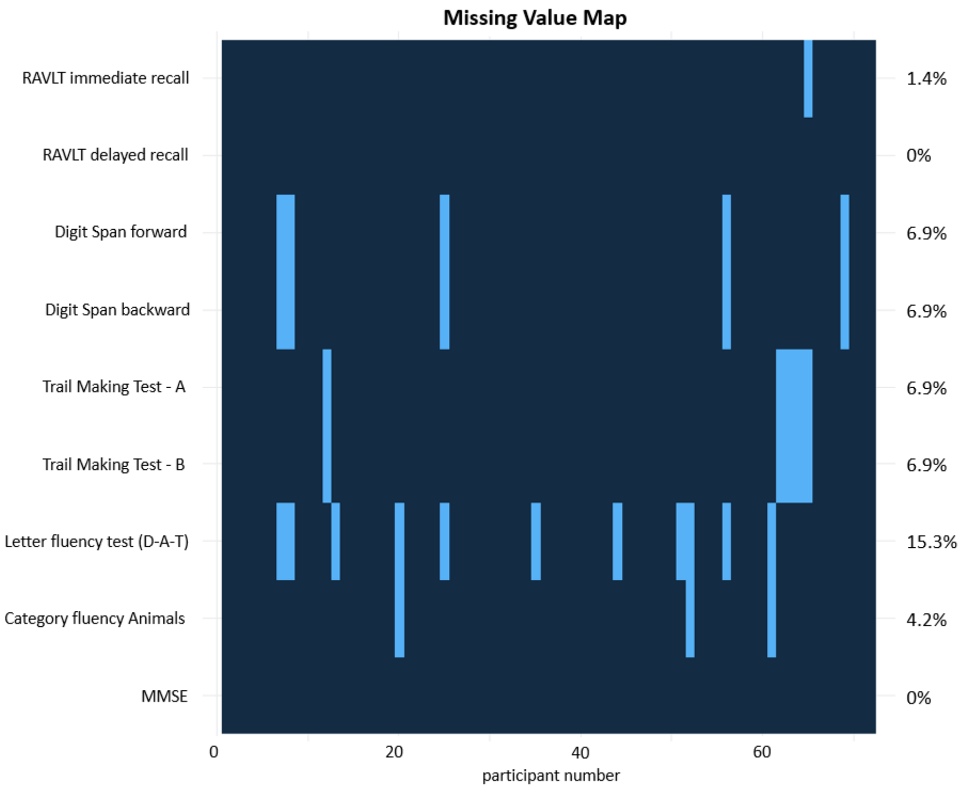


**sFigure-1. Map of missing values prior to imputation in the cognition-subsample.**

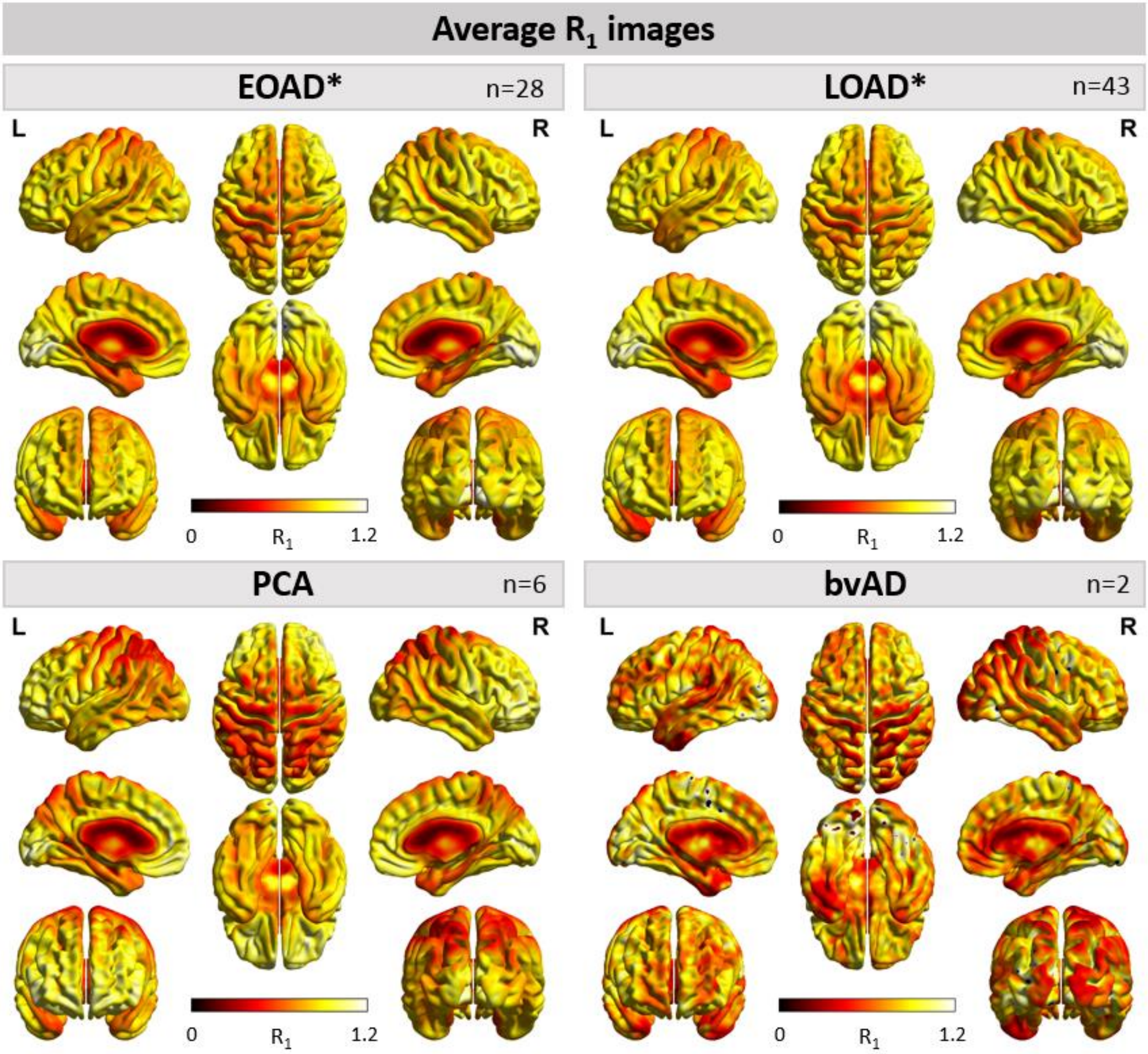


**sFigure-2. Average [$^{18}$F]flortaucipir $R_1$ images for early- and late-onset AD, PCA and bvAD.** Average images of all early-onset Alzheimer's disease (EOAD), late-onset AD (LOAD), posterior cortical atrophy (PCA) patients and behavioral variant AD (bvAD) patients on a scale ranging from $R_1$ 0 to 1.2. Low $R_1$ values represent low relative cerebral blood flow.

* Excluding atypical variants, posterior cortical atrophy (PCA) patients and behavioral variant AD (bvAD) patients.

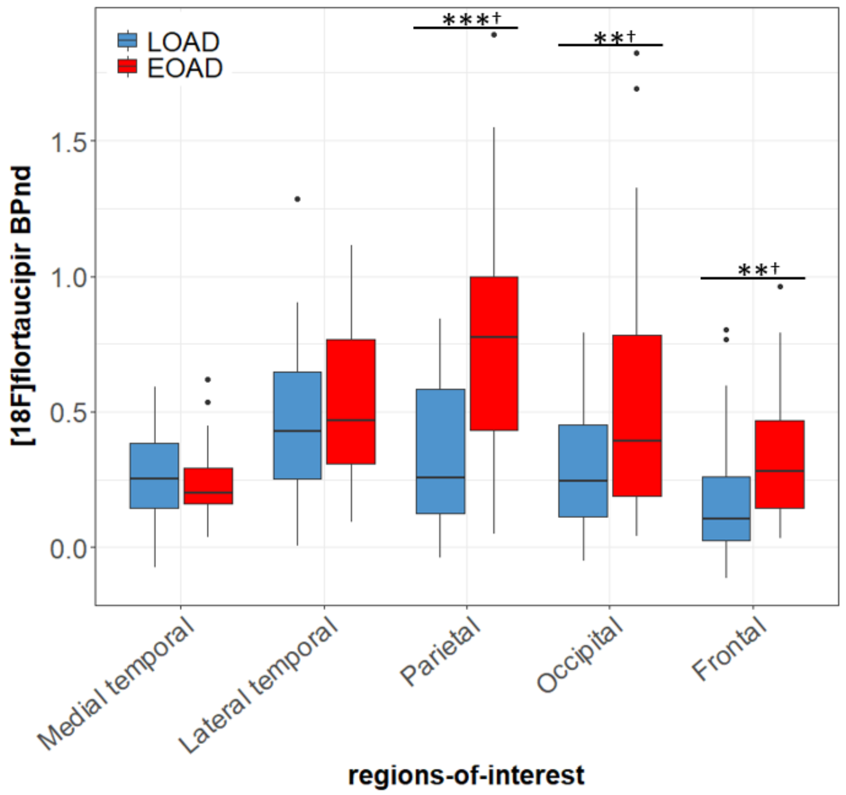


**sFigure-3. [$^{18}$F]flortaucipir BP$_{ND}$ for all subjects, excluding atypical (6 PCA and 2 bvAD) AD cases.** Differences between early-onset (EOAD) and late-onset AD (LOAD) were assessed using ANOVA, adjusted for sex. *$p<0.05$, **$p<0.01$, ***$p<0.001$, †$p_{FDR}<0.05$.

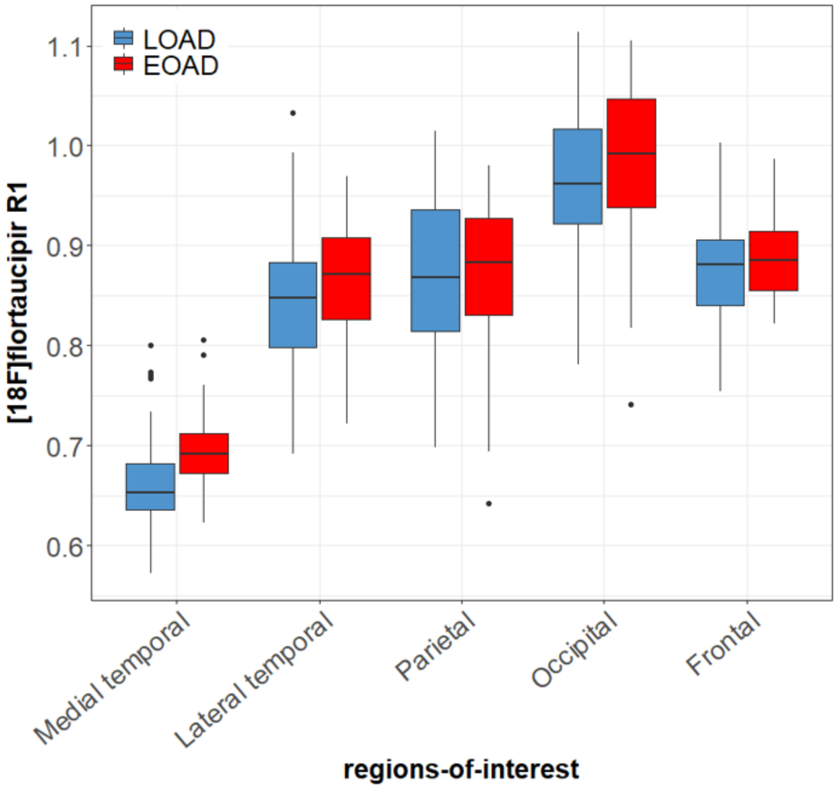


**sFigure-4. [$^{18}$F]flortaucipir $R_1$ for all subjects, excluding atypical (6 PCA and 2 bvAD) AD cases.** Differences between early-onset (EOAD) and late-onset AD (LOAD) were assessed using ANOVA, adjusted for sex.
